# Retracted randomized trials from super-retractors and top-cited scientists with multiple retractions

**DOI:** 10.1101/2025.11.23.25340834

**Authors:** Chunwei Lyu, Minoo Matbouriahi, Florian Naudet, John P.A. Ioannidis, Ioana Alina Cristea

## Abstract

**Importance:** Multiple retractions from the same author often uncover issues affecting their entire work, such as having systematically altered or fabricated data.

**Objectives:** Evaluate the contribution of authors with most retractions (“super-retractors”) and top-cited scientists with multiple retractions to the retracted clinical trial literature.

**Design:** Retrospective cohort study, linking an openly available cohort (VITALITY) of 1330 retracted randomized clinical trials (RCTs) to three lists of scientists: super-retractors, totaling most retractions in the Retraction Watch Leaderboard, and top-cited scientists, over the entire career or in the most recent single year, who accumulated 10 or more retractions not due to editor/publisher errors. The VITALITY cohort was updated up to November 2024. The three author lists were updated in August 2025.

**Participants:** 30 super-retractors, 163 career-long and 174 single-year scientists totaling 10 or more retractions.

**Main outcomes:** Authorship and characteristics of retracted RCTs (publication and retraction year, time between publication and retraction, number of citations).

**Results:** 6/30 super-retractors, representing Anesthesiology and Endocrinology & Metabolism, co-authored 290/1330 retracted RCTs (22%). 18/163 career-long top-cited scientists with at least 10 retractions, representing 10 fields, co-authored 327/1330 trials (25%), 275 (84%) of which were also co-authored by a super-retractor. 7/174 single-year top-cited scientists with at least 10 retractions co-authored 50 retracted trials; all of them were also among the career-long top-cited scientists with at least 10 retractions. Articles with super-retractors authors vs not were published earlier (median (IQR)= 2000 (1997-2005) vs 2020 (2014-2022)); retracted earlier (median (IQR)= 2013 (2012-2019) vs 2023 (2018.5-2023)); had a longer lag between publication and retraction, (median (IQR)= 5111 (3560-6820) vs 482 (330-1119) days); and accrued more citations (median (IQR)= 21 (12-42) vs 5 (1-19)). In multivariable regression models, only time to retraction (β = 0.02, *P* < 0.001) was significantly and positively associated with total citations. Results were similar when comparing retracted articles from top-cited scientists with at least 10 retractions versus other articles.

**Conclusions and relevance:** In this cohort study of 1330 retracted RCTs, a small number of influential authors, often co-authors and concentrated across few fields of medicine and countries, account for a significant proportion of retracted clinical trials.

**Key points:** *Question:* What is the contribution of the authors with most retractions (“super-retractors”) and of those top-cited with multiple retractions to the retracted randomized clinical trials literature?

*Findings:* In this cohort study, six super-retractors, from Anesthesiology and Endocrinology & Metabolism, co-authored one fifth of all retracted trials, while 18 top-cited scientists with over 10 retractions co-authored a quarter of them. Articles co-authored by super-retractors or by top-cited scientists with multiple retractions were published and retracted earlier, took longer to retract and accumulated more citations.

*Meaning:* Retracted clinical trials are disproportionately associated with a small number of influential authors, often co-authors and concentrated across few subfields of medicine and countries.

## Introduction

Retracted randomized controlled trials (RCTs) have an enduring and multiplicative impact of the evidence ecosystem, by distorting findings of systematic reviews and meta-analyses^1,2^, which then percolate into clinical practice guidelines^1^. Concerningly, only a part of RCTs that would qualify for a retraction end up retracted^3^. According to the guidelines by the Committee on Publication Ethics (COPE)^4^, “clear evidence of major irregularities in the data or images (…) that compromise the reliability of the findings” would warrant retraction. By this metric, “zombie” RCTs^5^, i.e. that appear to be fake or where data blatantly lacks credibility, should all have been either rejected by journals or retracted^6^. This does not seem to be happening. For example, employing the INSPECT-SR tool for assessing trustworthiness of 95 RCTs included in Cochrane systematic reviews, 24 (25%) and 6 (6%) were identified as having some and respectively serious concerns^7^. Only 2 of these flagged trials had been retracted or accompanied by expressions of concern.

Previous research has described super-retractors, scientists who have amassed numerous retractions^8,9^. Similarly, recent work examined influential, highly-cited scientists with retractions not due to publisher/journal error^10^, a subset of whom had over10 retractions. Multiple retractions from the same author often stimulate investigations that uncover issues affecting an author’s entire work, such as having systematically altered or fabricated data. However, there are no estimations of the contribution of super-retractors or top-cited authors with multiple retractions to the corpus of retracted RCTs. We aimed to quantify this contribution, based on a previously published and openly available complete cohort of retracted RCTs^1^ (VITALITY).

## Methods

### Ethical approval

Ethical approval was not requested for this study because it involved data extracted from published research and the publicly available datasets on which this research was based. No human participants were involved.

### Study population

The study is a retrospective cohort study and STROBE reporting guidelines^11^ were followed. The VITALITY cohort^1^ includes 1330 RCTs with notification of retraction, identified from the Retraction Watch (RW) (updated November 5^th^ 2024). To assemble the VITALITY cohort, records labelled as “Clinical Study” or as “Research articles” limited to subjects of “(HSC) Health Sciences” were retrieved from RW. Subsequently, two independent researchers reviewed titles, abstracts and full-texts and selected all studies that reported the design as a randomized trial in the methods. Multiple analyses from the same primary RCT were retained only if data were analyzed as per original randomization. No restrictions were placed for reasons for retraction or other characteristics.

References and study characteristics for the cohort are openly available (https://osf.io/7eazq/). We linked the VITALITY cohort with three groups of authors with multiple retractions.

Group 1 came from the RW Leaderboard (https://retractionwatch.com/the-retraction-watch-leaderboard/), August 26^th^ 2025 update, and includes 30 researchers with highest number of retractions (range, 33 to 221).

The two other author groups came from the science-wide author databases of standardized citation indicators (https://elsevier.digitalcommonsdata.com/datasets/btchxktzyw/8), August 2025 update: scientists that are top-cited over their career (career-long, Group 2) and in 2024 (single recent-year, Group 3). Top-cited was defined^12^ as being among the 100000 scientists or in the top 2% percentile among authors in the same subfield by a composite of standardized values of citation indicators.^13^. In the science-wide author databases, retractions are only counted if not due to publisher/journal error and if the paper was not replaced by a revised version. We selected scientists from the top-cited career-long and respectively single-year cohorts who amassed at least 10 such retractions (n=163 scientists in Group 2, n=174 in Group 3).

### Data extraction

References in the publicly available VITALITY dataset include truncated authors lists, so one author (CL) extracted the full reference that included full names of all authors for each retracted RCT, using the available DOI. Subsequently, for each complete reference, two researchers (CL and MM) independently checked each of the authors names against the lists of scientists in Groups 1, 2 and 3 using the Excel “search” function and manually examining each hit. For each article, the researchers noted whether any authors from Groups 1, 2 or 3 were involved as authors.

From the VITALITY dataset, we compiled characteristics of retracted trials, namely publication and retraction dates (month-date-year), from which we calculated the time (days) from publication to retraction. Additionally, one researcher (CL) independently extracted number of citations from Scopus as of November 21, 2025.

### Data analysis

Inter-rater agreement was assessed with the kappa statistic, separately for the three Groups of authors (super-retractors, top-cited career-long and single-year).

Results are presented descriptively as counts, proportions and confidence intervals (CIs) calculated with the binomial exact method, and medians and interquartile ranges (IQR). As we expected high level of correlation between the variables considered, we conducted Pearson correlation with *P*-values corrected for multiple comparisons with the Sidak correction. We compared publication and retraction year, time from publication to retraction and total citations between groups (i.e. involving super-retractors/top-cited career-long/top-cited single-year versus not) with Wilcoxon rank-sum test. To disentangle the effects on citation counts of authorship from super-retractors or top-cited scientists with multiple retractions from survival effects (i.e., articles that survive longer in the literature are cited more), we conducted multivariable linear regression analysis including presence of super-retractor/top-cited author as binary variable, time between retraction and publication (days) as continuous and their interaction. Significant interaction effects were followed up by univariate regression separate for papers involving super-retractors/top-cited scientists or not.

Analyses were conducted in Stata SE 19.0 and Claude Sonnet 4.5 was used for checking code and for data visualization.

## Results

Fifteen/163 scientists in Group 2 and 8/174 in Group 3 were also super-retractors (Group 1). Kappa inter-rater agreement was 100% for scientists in Group 1 and respectively Group 2, and 99.92% (1 discrepancy) for Group 3.

Of 30 super-retractors, six (Table 1) had co-authored 290 of the 1330 retracted RCTs (22%). 52/290 (18%) papers had been co-authored by more than one super-retractor. The 6 super-retractors belonged to two subfields (Anesthesiology (n=3) and Endocrinology & Metabolism (n=3)) and all but one were affiliated with Japanese institutions. Retracted RCTs represented between 3% and 79% of all their retracted articles. For 3 super-retractors, over half of their retracted papers were RCTs.

**Table 1.** Retracted randomized trials authored by known super-retractors.

| Super-retractor | Subfield | Country | RCTs as 1<sup>st</sup> author | RCTs as co-author | Total retracted RCTs | Total retracted articles<sup>b</sup> | % retracted RCTs vs articles (95% CI) |
| --- | --- | --- | --- | --- | --- | --- | --- |
| Joachim Boldt | Anesthesiology | DE | 72 | 47 | 119 | 221 | 54 (47– 61) |
| Yoshitaka Fujii | Anesthesiology | JP | 111 | 10 | 121 | 172 | 70 (63 –77) |
| Yoshihiro Sato | Endocrinology & Metabolism | JP | 26 | 4 | 30 | 124 | 24 (17 – 33) |
| Hironobu Ueshima | Anesthesiology | JP | 3 | 1 | 4 | 124 | 3 (1 – 8) |
| Jun Iwamoto | Endocrinology & Metabolism | JP | 5 | 19 | 24 | 91 | 26 (18 – 37) |
| Yuhji Saitoh | Anesthesiology | JP | 20 | 24 | 44 | 56 | 79 (66 – 88) |

*Note.*
CI, confidence interval; DE, Germany; JP, Japan; RCT, randomized controlled trials; %, proportion
<sup>a</sup>According to the updated science-wide author databases of standardized citation indexes
<sup>b</sup>According to the Retraction Watch Database, list updated August 25<sup>th</sup> 2025

Eighteen of the 163 career-long top-cited scientists with at least 10 retractions (eTable 1) co-authored 327/1330 retracted trials (25%). Five of these authors were also super-retractors. The 18 top-cited scientists covered 10 subfields, but many concentrated on Anesthesiology (n=4 authors) and Cardiovascular System & Hematology (n=4). They represented 8 countries, with most in Japan (n=7) and the US (n=4). 7/18 had co-authored only a single retracted trial while for another 7 retracted RCTs represented half of their retracted papers. Overall, 275/327 retracted RCTs (84%) involved a super-retractor.

Seven out of 174 top-cited scientists in 2024 with at least 10 retractions (eTable 2) co-authored 50 /1330 retracted trials (4%). All 7 authors were also among the top-cited career-long. Three authors had co-authored a single retracted trial, while for 4 authors retracted RCTs represented over half of their retracted papers. None of the authors themselves was on the RW leaderboard, but 14 out of 50 (28%) retracted RCTs were co-authored with a super-retractor (Kei Saitoh).

As displayed in Table 2, retracted RCT articles with super-retractor authors were published earlier, median (IQR) of 2000 (1997-2005) vs 2020 (2014-2022) (p<.00001 by Wilcoxon rank-sum test); retracted earlier, median (IQR) of 2013 (2012-2019) vs 2023 (2018.5-2023) (p<.00001); had a higher lag between publication and retraction, median days (IQR) of 5111 (3560-6820) vs 482 (330-1119) (p<.00001); and amassed more citations, median (IQR) of 21 (12-42) vs 5 (1-19) (p<.00001), than retracted RCT articles without super-retractor authors. Similarly, retracted articles published by top-cited career-long scientists with at least 10 retractions were published earlier, median (IQR)= 2001 (1998-2007) versus 2021 (2014-2022) (p<.00001 by Wilcoxon rank-sum test); retracted earlier median (IQR)= 2014 (2012-2020) versus 2023 (2019-2023) (p<.00001); had a longer lag between retraction and publication, median days (IQR)= 4748 (2708-5872) vs 469 (325-1058) (p<.00001); and more citations, median (IQR)= 24 (14-47) vs 4 (1-16) (p<.00001) than other retracted RCT articles. Results were similar for articles published by single-year top-cited scientists versus other retracted RCT articles (Table 2). The results of correlation analyses are displayed in eTable 3. Total citations had small, but significant negative relationships with publication year (Pearson r = -0.144, *P* < 0.001), retraction year (Pearson r = -0.128, *P* < 0.001) and a similarly small, but significant positive relationship with time between publication and retraction (Pearson r = 0.097, *P* = 0.009). Correlations with the presence of authors from any of the three Groups were non-significant (all P-values> 0.002).

**Table 2.** Characteristics of retracted randomized trials involving super-retractors, career-long and 2024 top-cited scientists with at least 10 retractions versus trials not involving them.

|  | Article, Count (%) | Publication year, median (IQR) | P-value<sup>a</sup> | Retraction year, median (IQR) | P-value | Time (days) to retraction, median (IQR) | P-value | Citations, median (IQR)<sup>b,c</sup> | P-value |
| --- | --- | --- | --- | --- | --- | --- | --- | --- | --- |
| Super-retractor | 290 (22%) | 2000 (1997-2005) | <.00001 | 2013 (2012-2019) | <.00001 | 5111 (3560-6820) | <.00001 | 21 (12-42) | <.00001 |
| No super-retractor | 1040 (78%) | 2020 (2014-2022) |  | 2023 (2018.5-2023) |  | 482 (330-1119) |  | 5 (1-19) |  |
| Top-cited career-long $\geq 10$ retractions | 327 (25%) | 2001 (1998-2007) | <.00001 | 2014 (2012-2020) | <.00001 | 4748 (2708-5872) | <.00001 | 24 (14-47) | <.00001 |
| No top-cited career-long $\geq 10$ retractions | 1003 (75%) | 2021 (2014-2022) | | 2023 (2019-2023) | | 469 (325-1058) | | 4 (1-16) | |
| Top-cited 2024 $\geq 10$ retractions | 50 (4%) | 2010 (2005-2015) | <.00001 | 2017 (2016-2022) | 0.049 | 2557.5 (1597-4718) | <.00001 | 38 (18-65) | <.00001 |
| No top-cited 2024 $\geq 10$ retractions | 1280 (96%) | 2018 (2008-2022) | | 2022 (2015-2023) | | 611 (358-2504) | | 7 (2-24) | |
| Total | 1330 | 2017 (2007-2022) |  | 2022 (2015-2023) |  | 637.5 (362-2690) |  | 13 (4-42) |  |

Note. IQR, interquartile range
<sup>a</sup> Calculated with the Wilcoxon rank-sum test
<sup>b</sup> Citations counts were extracted from Scopus
<sup>c</sup> For 12 trials, number of citations was not available

In multivariable regression models (eTable 4, eResults, eFigure 1), time to retraction (β = 0.02, *P* < 0.001), but not presence of super-retractors (β = 24.7, *P* = 0.098) was significantly and positively associated with total citations. The interaction between time to retraction and presence of super-retractors was significant and negative (β = -0.02, *P* < 0.001). Similarly (eTable 4, eResults, eFigure 2), both time to retraction (β = 0.01, *P* < 0.001) and presence of top-cited career-long scientists with at least 10 retractions (β = 35.26, *P* = 0.007) were significantly and positively associated with number of citations, but their interaction had a significant and negative effect (β = -0.01, *P* < 0.001). For articles co-authored by scientists top-cited in 2024 with at least 10 retractions (eTable 4), only time to retraction was positively associated with citation counts (β = 0.004, *P* = 0.001), and there was no significant interaction effect.

## Discussion

Six super-retractors, representing two fields of medicine, account for one fifth of all retracted randomized trials. Similarly, 18 top-cited scientists with over 10 retractions account for a quarter of all retracted RCTs. All but 1 of the super-retractors involved in retracted RCTs were also among the top-cited over their career. There was a large overlap between retracted trials attributed to super-retractors and those attributed to career-long highly cited scientists who amassed at least 10 retractions. Retracted RCT articles co-authored by super-retractors or by top-cited scientists with multiple retractions were published and retracted earlier, took longer to retract and, in uncorrected analysis, accumulated more citations than retracted RCT articles not involving such authors. In correlation analysis, retraction- and publication year, as well as time to retraction showed a modest association with citation counts, but not author status. In multivariable analysis, time to retraction was consistently positively related to citations, as expected, while author status was no longer significantly related with citations. There was a significant interaction effect between time to retraction and author status: for articles by authors who were not super-retractors or top-cited over their career, citations gradually increased over time, while there was no such trend for articles attributed to super-retractors or top-cited career-long scientists.

Multiple retractions from the same author stimulate efforts to inspect other publications belonging to them, often leading to more retractions. For example, the work of super-retractors has been thoroughly scrutinized, with an analysis from 2019^14^ indicating that only 9% of the papers eligible for retraction remained not retracted. Conversely, the work of top-cited scientists with many retracted publications has received less attention; additional papers by these authors may deserve scrutiny. Nevertheless, the enhanced examination likely led to a substantial overrepresentation of publications from super-retractors or top-cited scientists with multiple retractions within the current population of retracted randomized trials. As probably all papers from super-retractors were examined and many subsequently retracted, the cohort of retracted trials included a mix of more consequential, and thus more cited papers, and more peripheral ones, which received significantly less attention from the scientific community. A similar phenomenon could have occurred to papers from top-cited career-long scientists, which largely overlapped with papers attributed to super-retractors. Additional scrutiny to top-cited scientists who currently have less than 10 retractions, so were not included in our analysis, might lead to future retractions and increase their representation within the cohort of retracted trials. Super-retractor papers get more citations probably mostly because they have had more time before they were retracted. However, some of the authors with retractions who are currently not super-retractors may eventually have additional retractions and may reach super-retractor status in the future. By then, it is possible that their late-retracted papers would have accumulated many more citations before being taken off the literature. In contrast, papers from researchers top-cited in 2024 were fewer and only time to retraction was associated with citation counts. Some of the authors who currently have few, sporadic retractions may end up accumulating more retractions in the future and they may also enter the group of super-retractors.

Retracted randomized trials belonging to super-retractors or scientists top-cited over their entire career were published earlier and took longer to retract, meaning there was more time to identify and investigate publications, formulate allegations and follow-up on retractions. For example, concerns about Yoshihiro Sato^9^, one of the most prolific super-retractors, had been raised almost 10 years before the first retractions occurred, in letters to the editor, and systematic investigation of his work by other researchers took almost 3 years to get published^15^ and for the first retractions to begin. Moreover, investigation of multiple papers from the same author often attracts examination of co-authors^9^, which in the case of Sato, led to another super-retractor in his frequent co-author, Jun Iwamoto. Another frequent co-author, Kei Satoh, was only included on the top-cited career-long list in the current analysis. Co-authorship patterns at least partly account for the overrepresentation of some countries in our cohort of retracted RCTs.

Beyond investigation of co-authors of prolific retractors, some fields, most notably Anesthesia, have been extensively scrutinized, also leading to overrepresentation in our sample. An analysis^16^ of anesthesia articles retracted until 2017 identified 350 retracted articles, with 4 individuals responsible to almost 60% of all retracted articles. Though these articles were not restricted to randomized trials, two of the four authors identified (Yoshitaka Fujii and Joachim Boldt) were also on our list of super-retractors and top-cited career-long authors of RCTs and another (Scott S. Reuben) only on the top-cited list. These authors worked independently on different topics, so their large, combined output of retracted RCTs cannot be attributed to co-authorship.

Notably, several authors with many retractions were top-cited for their career-long citation impact, but only a minority of them continued to be top-cited in the most recent single year. This may reflect reputational loss and decline in citations to their publication corpus. An earlier analysis^17^ evidenced that authors who had a retraction experienced a citation penalty of 10% (20% if the retraction was related to misconduct) for their non-retracted work. This penalty is likely more severe for authors with multiple retractions and might extend to their frequent co-authors. Concentration of prolific retractors in some subfields and countries may reflect either systemic problems in these subfields and countries or better sensitization towards detecting fraud. Conversely, it is possible that the overrepresentation of some fields and countries masks insufficient scrutiny of other fields or inadequate detection methods in other countries.

Our study has several limitations. First, retractions represent only a fraction of the literature should be retracted, following guidelines such as COPE^4^. Retracted trials, even when clearly linked to data fabrication, fraud or severe methodological problems, are not randomly selected sample of zombie trials^5^, which limits drawing inferences about factors associated with misconduct or inadequate research practices from cohort of retracted trials. Some authors with few retracted papers may accumulate more retractions in the future and some fields are receiving heightened scrutiny, such as obstetrics and gynecology^18,19^, potentially changing the profiles emerging in our current data. Recent proposals^19^ to examine the collected body of evidence coming from an author or author group suspected of misconduct might uncover other super-retractors. Second, the VITALITY cohort includes trials retracted for any reason, so also for problems not related to data fabrication or severe flaws, thus precluding the use of retraction as a proxy for misconduct or fraud. However, all three lists of authors including researchers who accumulated multiple retractions for reasons not imputable to publisher or journal error, thereby increasing the likelihood that their accumulated retractions reflect at least inadequate research practices, if not overt misconduct. Third, analysis of citations is confounded by the fact retracted papers continue to be cited after retraction^20^, even when retraction was explicitly linked to misconduct^21^, and we did not distinguish citations before or after the retraction. Fourth, the dates of update of the three cohorts included are slightly different, with VITALITY dataset updated in November 2024 and the three author groups in August 2025. Moreover, citations counts for VITALITY cohort were extracted de novo in November 2025, though this update is unlikely to have produced a major shift in citations.

The disproportionate role of super-retractors and top-cited authors with multiple retractions, their interconnections as expressed by co-authorships, and the narrow concentration across few fields of medicine and countries may provide useful leads to help unravel fraudulent literature at scale.

## Supporting information

STROBE checklist

## Data Availability

The VITALITY cohort (https://osf.io/7eazq/), the science-wide author database of standardized citation indexes ((https://elsevier.digitalcommonsdata.com/datasets/btchxktzyw/8) and the Retraction Watch Leaderboard (https://retractionwatch.com/the-retraction-watch-leaderboard/) are publicly available. The linked datasets and additional extracted variables, as well as STATA code to reproduce analysis are publicly available on Zenodo after publication (Version v210.5281/zenodo.1863054).

https://zenodo.org/records/18630545

## Conflict of Interest Disclosures

No conflicts of interest.

## Funding

Chunwei Lyu is funded by the China Scholarship Council (award number 202410710001). Minoo Matbouriahi is supported by the doctoral network MSCA-DN SHARE-CTD (HORIZON-MSCA-2022-DN-01, grant agreement 101120360), funded by the EU. Florian Naudet received funding from the French National Research Agency, the French Ministry of Health, and the French Ministry of Research and is a work package leader in the OSIRIS project (Open Science to Increase Reproducibility in Science, grant agreement 101094725) and for the doctoral network MSCA-DN SHARE-CTD (HORIZON-MSCA-2022-DN-01, grant agreement 101120360), funded by the EU. John P. A. Ioannidis is supported by an unrestricted gift from Sue and Bob O’Donnell to Stanford University. Ioana Alina Cristea is supported by a European Research Council (ERC) Starting Grant DECOMPOSE (grant agreement: 101042701, https://cordis.europa.eu/project/id/101042701), funded by the European Union (EU) and by the doctoral network MSCA-DN SHARE-CTD (HORIZON-MSCA-2022-DN-01, grant agreement 101120360), funded by the EU.

### Role of the Funder/Sponsor

The funders had no role in the study design, data collection and analysis, decision to publish, or preparation of the manuscript.

### Access to data and data analysis

IAC had full access to all the data in the study and takes responsibility for the integrity of the data and the accuracy of the data analysis.

## Author contributions

Conceptualization: IAC, JPAI; Data curation: CL, MM; Formal analysis: IAC; Methodology: IAC, FN, JPAI; Funding acquisition: All; Supervision: FN, JPAI, IAC; Writing-original draft: IAC, JPAI; Writing-review and editing: All

## Notes

### Competing Interest Statement

The authors have declared no competing interest.

### Summary of Updates

This version was revised following peer-review.

## References

1. Xu C, Fan S, Tian Y, et al. Investigating the impact of trial retractions on the healthcare evidence ecosystem (VITALITY Study I): retrospective cohort study. BMJ. 2025;389:e082068. doi:10.1136/bmj-2024-082068

2. Graña Possamai C, Cabanac G, Perrodeau E, Ghosn L, Ravaud P, Boutron I. Inclusion of Retracted Studies in Systematic Reviews and Meta-Analyses of Interventions: A Systematic Review and Meta-Analysis. JAMA Intern Med. Jun 1 2025;185(6):702–709. doi:10.1001/jamainternmed.2025.0256

3. Van Noorden R. Medicine is plagued by untrustworthy clinical trials. How many studies are faked or flawed? Nature 2023. p. 454–458.

4. COPE Council. COPE Guidelines: Retraction Guidelines. August 2025. 08/2025. doi:10.24318/cope.2019.1.4 Accessed 02/06/2026. https://publicationethics.org/guidance/guideline/retraction-guidelines

5. Ioannidis JPA. Hundreds of thousands of zombie randomised trials circulate among us. Anaesthesia. 2021;76(4):444–447. doi:10.1111/anae.15297

6. Carlisle JB. False individual patient data and zombie randomised controlled trials submitted to Anaesthesia. Anaesthesia. 2021;76(4):472–479. doi:10.1111/anae.15263

7. Wilkinson J, Heal C, Antoniou GA, et al. Assessing the feasibility and impact of clinical trial trustworthiness checks via an application to Cochrane Reviews: Stage 2 of the INSPECT-SR project. Journal of Clinical Epidemiology. 2025/08/01/ 2025;184:111824. doi:10.1016/j.jclinepi.2025.111824

8. Wise J. Boldt: the great pretender. BMJ : British Medical Journal. 2013;346:f1738. doi:10.1136/bmj.f1738

9. Kupferschmidt K. Researcher at the center of an epic fraud remains an enigma to those who exposed him. Science: Science; 2018.

10. Ioannidis JPA, Pezzullo AM, Cristiano A, Boccia S, Baas J. Linking citation and retraction data reveals the demographics of scientific retractions among highly cited authors. PLOS Biology. 2025;23(1):e3002999. doi:10.1371/journal.pbio.3002999

11. von Elm E, Altman DG, Egger M, Pocock SJ, Gøtzsche PC, Vandenbroucke JP. The Strengthening the Reporting of Observational Studies in Epidemiology (STROBE) statement: guidelines for reporting observational studies. Ann Intern Med. Oct 16 2007;147(8):573–7. doi:10.7326/0003-4819-147-8-200710160-00010

12. Ioannidis JPA, Baas J, Klavans R, Boyack KW. A standardized citation metrics author database annotated for scientific field. PLOS Biology. 2019;17(8):e3000384. doi:10.1371/journal.pbio.3000384

13. Ioannidis JPA, Klavans R, Boyack KW. Multiple Citation Indicators and Their Composite across Scientific Disciplines. PLOS Biology. 2016;14(7):e1002501. doi:10.1371/journal.pbio.1002501

14. McHugh UM, Yentis SM. An analysis of retractions of papers authored by Scott Reuben, Joachim Boldt and Yoshitaka Fujii. Anaesthesia. 2019;74(1):17–21. doi:10.1111/anae.14414

15. Bolland MJ, Avenell A, Gamble GD, Grey A. Systematic review and statistical analysis of the integrity of 33 randomized controlled trials. Neurology. Dec 6 2016;87(23):2391–2402. doi:10.1212/wnl.0000000000003387

16. Nair S, Yean C, Yoo J, Leff J, Delphin E, Adams DC. Reasons for article retraction in anesthesiology: a comprehensive analysis. Can J Anaesth. Jan 2020;67(1):57–63. Raisons justifiant la rétractation d’un article en anesthésiologie: une analyse exhaustive. doi:10.1007/s12630-019-01508-3

17. Azoulay P, Bonatti A, Krieger JL. The career effects of scandal: Evidence from scientific retractions. Research Policy. 2017/11/01/ 2017;46(9):1552–1569. doi:10.1016/j.respol.2017.07.003

18. Patabendige M, Rolnik DL, Li W, Mol BW, Li W. Data-sharing and trustworthiness of trials evaluating cervical ripening in induction of labour: a meta-epidemiological study of randomised controlled trials. eClinicalMedicine. 2025;85 doi:10.1016/j.eclinm.2025.103346

19. Nielsen J, Bordewijk EM, Gurrin LC, et al. Assessing the scientific integrity of the collected work of one author or author group. J Clin Epidemiol. Apr 2025;180:111603. doi:10.1016/j.jclinepi.2024.111603

20. Woo S, Walsh JP. On the shoulders of fallen giants: What do references to retracted research tell us about citation behaviors? Quantitative Science Studies. 2024;5(1):1–30. doi:10.1162/qss_a_00303

21. Candal-Pedreira C, Ruano-Ravina A, Fernández E, Ramos J, Campos-Varela I, Pérez-Ríos M. Does retraction after misconduct have an impact on citations? A pre-post study. BMJ Global Health. 2020;5(11):e003719. doi:10.1136/bmjgh-2020-003719

