## Supplementary material for "Retracted randomized trials from super-retractors and top-cited scientists with multiple retractions"

### **eTable 1. Retracted randomized trials authored by career-long top-cited scientists with at least 10 retractions**

| **Top-cited (career-long) & ≥ 10 retractions** | **Subfield^a^** | **Country** | **RCTs as 1^st^ author** | **RCTs as**  **co-author** | **Total retracted RCTs** | **Total retracted articles^i^** | **% retracted RCTs vs articles (95% CI)** |
| --- | --- | --- | --- | --- | --- | --- | --- |
| Yoshitaka Fujii^b^ | Anesthesiology | JP | 111 | 10 | 121 | 168 | 72 (65 – 79) |
| Joachim Boldt^b^ | Anesthesiology | DE | 72 | 47 | 119 | 217 | 55 (48 – 62) |
| Yoshihiro Sato^b^ | Endocrinology & Metabolism | JP | 26 | 4 | 30 | 119 | 25 (18 – 34) |
| Jun Iwamoto^b^ | Endocrinology & Metabolism | JP | 5 | 19 | 24 | 89 | 27 (18 – 37) |
| Kei Satoh | Immunology | JP | 0 | 14 | 14 | 28 | 50 (31 – 69) |
| Scott S. Reuben | Anesthesiology | US | 14 | 0 | 14 | 17 | 82 (57 – 96) |
| Zatollah Asemi | Nutrition & Dietetics | IR | 3 | 9 | 12 | 13 | 92 (64 – 100) |
| Mohammad Reza Safarinejad | Urology & Nephrology | IR | 10 | 1 | 11 | 17 | 65 (38 – 86) |
| Giuseppe Derosa | Cardiovascular System & Hematology | IT | 10 | 0 | 10 | 10 | 100 (69 – 100) |
| Hironobu Ueshima^ii^ | Anesthesiology | JP | 3 | 1 | 4 | 120 | 3 (0.9 – 8) |
| Hideo Matsumoto | Orthopedics | JP | 0 | 4 | 4 | 26 | 15 (4 – 35) |
| H.J. Eysenck | Social Psychology | UK | 0 | 1 | 1 | 20 | 5 (0.1 – 25) |
| Piero Anversa | Cardiovascular System & Hematology | US | 0 | 1 | 1 | 19 | 5 (0.1 – 26) |
| Annarosa Leri | Cardiovascular System & Hematology | US | 0 | 1 | 1 | 19 | 5 (0.1 – 26) |
| Jan Kajstura | Cardiovascular System &  Hematology | US | 0 | 1 | 1 | 18 | 5 (0.1 – 27) |
| Walid Kamal Abdelbasset | Food Science | EG | 1 | 0 | 1 | 17 | 6 (0.1 – 29) |
| Mitsuhiro Osame | Neurology & Neurosurgery | JP | 0 | 1 | 1 | 14 | 7 (0.1 – 34) |
| Bruno Vellas | Neurology & Neurosurgery | FR | 0 | 1 | 1 | 10 | 1 (0.2 – 45) |

*Note.*

CI, confidence interval; RCT, randomized controlled trials, DE, Germany; EG, Egypt; FR, France; IT, Italy; IR, Iran; JP, Japan; UK, United Kingdom; US, United States

^a^ According to the updated science-wide author databases of standardized citation indexes

^b^ Also included on the Retraction Watch leaderboard

### **eTable 2. Retracted randomized trials authored by 2024 top-cited scientists with at least 10 retractions**

| **Top-cited (single year-long) & ≥ 10 retractions** | **Subfield^a^** | **Country** | **RCTs as 1^st^ author** | **RCTs as**  **co-author** | **Total retracted RCTs** | **Total retracted articles^i^** | **% retracted RCTs vs articles (95% CI)** |
| --- | --- | --- | --- | --- | --- | --- | --- |
| Kei Satoh | Immunology | JP | 0 | 14 | 14 | 28 | 50 (31 – 69) |
| Zatollah Asemi | Nutrition & Dietetics | IR | 3 | 9 | 12 | 13 | 92 (64 – 100) |
| Mohammad Reza Safarinejad | Urology & Nephrology | IR | 10 | 1 | 11 | 17 | 65 (38 – 86) |
| Giuseppe Derosa | Cardiovascular System & Hematology | IT | 10 | 0 | 10 | 10 | 100 (69 – 100) |
| H.J. Eysenck | Social Psychology | UK | 0 | 1 | 1 | 20 | 5 (0.1 – 25) |
| Walid Kamal Abdelbasset | Food Science | EG | 1 | 0 | 1 | 17 | 6 (0.1 – 29) |
| Bruno Vellas | Neurology & Neurosurgery | FR | 0 | 1 | 1 | 10 | 1 (0.2 – 45) |

*Note.*

CI, confidence interval; RCT, randomized controlled trials; EG, Egypt; FR, France; IT, Italy; IR, Iran; JP, Japan; UK, United Kingdom

^a^ According to the updated science-wide author databases of standardized citation indexes

### **eTable 3. Correlation between the main variables extracted**

| **Variable R (P-value)** | **1** | **2** | **3** | **4** | **5** | **6** | **7** |
| --- | --- | --- | --- | --- | --- | --- | --- |
| 1. Super-retractor (y/n) | 1.00 | 0.861ᵃ  (<0.001) | 0.03  (0.99) | -0.757ᵃ  (<0.001) | -0.367ᵃ  (<0.001) | 0.724ᵃ  (<0.001) | 0.030  (0.99) |
| 2. Top-cited career-long ≥ 10 retractions (y/n) |  | 1.00 | 0.346ᵃ  (<0.001) | -0.729ᵃ  (<0.001) | -0.403ᵃ  (<0.001) | 0.659ᵃ  (<0.001) | 0.07  (0.21) |
| 3. Top-cited in 2024 ≥ 10 retractions (y/n) |  |  | 1.00 | -0.082  (0.058) | -0.019  (>0.99) | 0.092ᵃ  (0.016) | 0.055  (0.63) |
| 4. Publication Year |  |  |  | 1.00 | 0.653ᵃ  (<0.001) | -0.835ᵃ  (<0.001) | -0.144ᵃ  (<0.001) |
| 5. Retraction Year |  |  |  |  | 1.00 | -0.132ᵃ  (<0.001) | -0.128ᵃ  (<0.001) |
| 6. Time to Retraction (days) |  |  |  |  |  | 1.00 | 0.097ᵃ  (0.009) |
| 7. Citations (Scopus)^a^ |  |  |  |  |  |  | 1.00 |

*Note.* Values shown are Pearson correlation coefficients.

^a^ N=1318

^b^ Statistically significant after Sidak correction for multiple comparisons (adjusted α = 0.0024 for 21 comparisons).

### **eTable4. Multivariable linear regression analysis with total citations (Scopus) as dependent variable**

| **Model** | **Variable** | **Coefficient (SE)** | ***P*-value** | **Overall *P*-value** | **R^2^** |
| --- | --- | --- | --- | --- | --- |
| 1 | Intercept | 7.13 (4.52) | 0.115 | <.001 | 0.03 |
|  | Time to retraction (days) | 0.02 (0.003) | <.001 |  |  |
|  | Super-retractor present (versus not)^a^ | 24.7 (14.93) | 0.098 |  |  |
|  | Time to retraction * Super-retractor interaction | -0.02 (0.004) | <.001 |  |  |
| 2 | Intercept | 10.01 (4.48) | 0.026 | <.001 | 0.03 |
|  | Time to retraction (days) | 0.01 (0.002) | <.001 |  |  |
|  | Top-cited career-long ≥ 10 retractions present (versus not)^b^ | 35.26 (12.98) | 0.007 |  |  |
|  | Time to retraction * Top-cited career-long ≥ 10 retractions interaction | -0.01 (0.003) | <.001 |  |  |
| 3 | Intercept | 17.14 | <.001 | 0.002 | 0.01 |
|  | Time to retraction (days) | 0.004 | 0.001 |  |  |
|  | Top-cited 2024 ≥ 10 retractions present (versus not)^c^ | 18.71 | 0.524 |  |  |
|  | Time to retraction *Top-cited 2024 ≥ 10 retractions interaction | 0.003 | 0.701 |  |  |

*Note.* Multivariable analysis included 1318 articles for which we could retrieve citation data.

^a^ 290 articles with a co-author super-retractor versus 1028 with no such co-author

^b^ 327 articles with a co-author top-cited career-long versus 991 with no such co-author

^c^ 50 articles with a co-author top-cited single-year versus 1268 with no such co-author

### **eResults**

Univariate regression analyses stratified by presence of super-retractors (eFigure 1) indicated that their papers versus those with no super-retractors had more baseline citations (intercept (SE)= 31.83 (4.70) versus 7.13 (5.06)), but a slower citation rate over time (slope= 0.0002 citations/day, *P* =0.807 vs 0.02 citations/day, *P* < 0.001). Univariate regression analyses (eFigure 2) similarly indicated that papers with top-cited career-long scientists co-authors versus those without them had more citations at baseline (intercept (SE)= 45.27 (7.93) versus 10.01 (4.89)), but average citation counts remained stable over time (slope= -0.001 citations/day, *P*=0.468 vs 0.01 citations/day, *P* < 0.001).

### **eFigure 1. Mean citations (±95% confidence interval) by survival time and super-retractor status in 200-day bins (≥5 papers)**

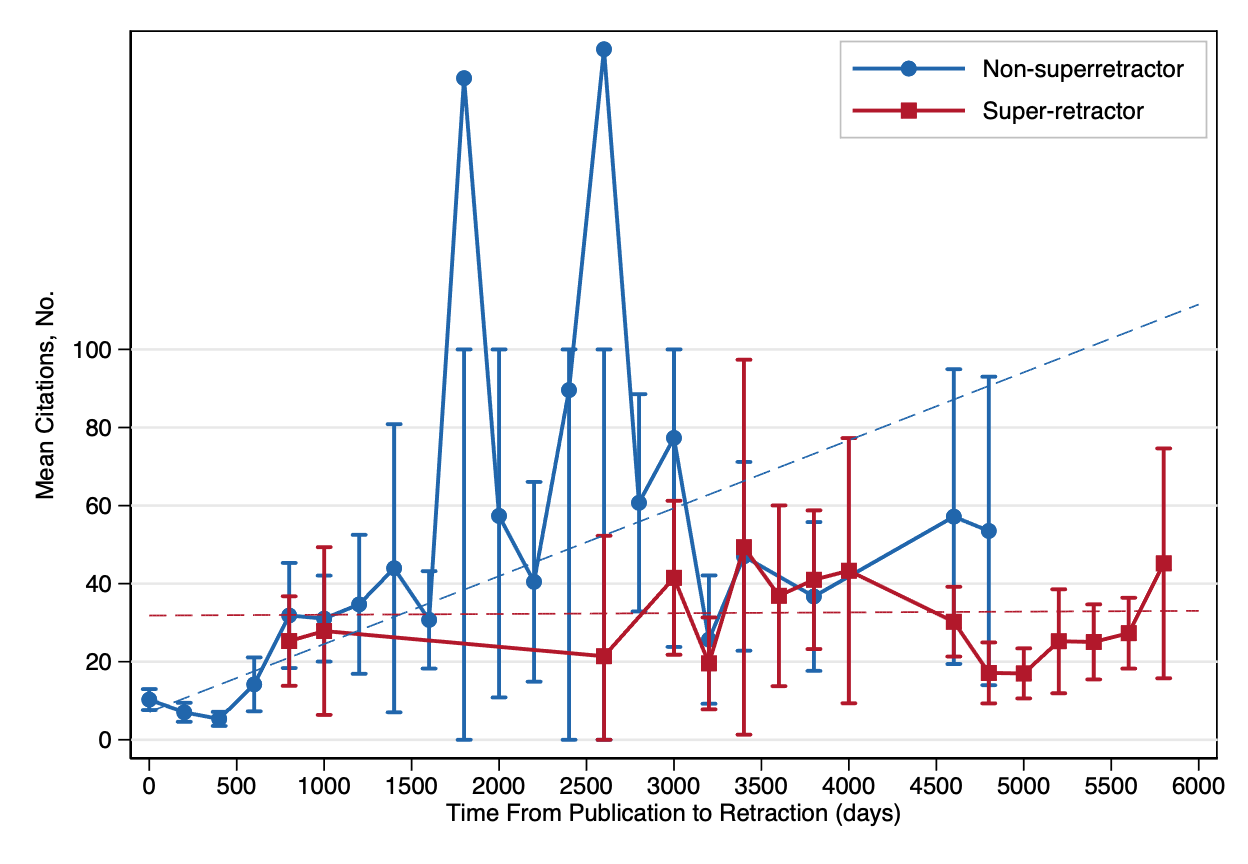

Dashed lines show model predictions. Papers not co-authored by super-retractors (blue circles) showed citation growth (0.017/day, P<.001); super-retractor papers (red squares) did not (0.0002/day, P=.81).

Data restricted to papers with a time-lag of ≤6,000 days between publication and retraction (97 papers) and bins with ≥5 papers (64 papers), leading to 1157 of 1318 papers (88%) plotted.

Confidence intervals capped at 100 for display.

### **eFigure 2. Mean citations (±95% confidence interval) by survival time and top-cited career-long status in 200-day bins (≥5 papers)**

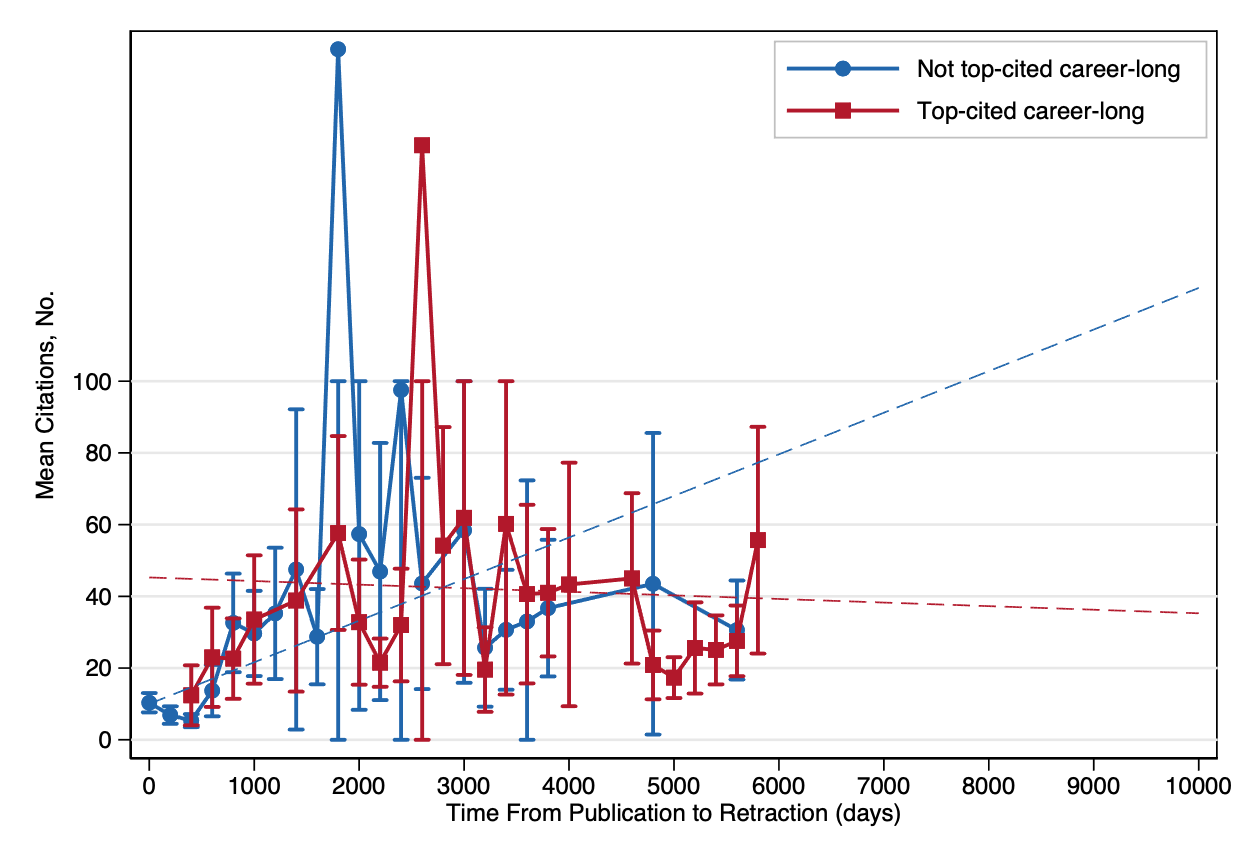

Dashed lines show model predictions. Papers not co-authored by top-cited career-long scientists (blue circles) showed citation growth (0.012/day, P<.001); papers by top-cited career-long authors (red squares) did not (-0.001/day, P=.47).

Data restricted to papers with a time-lag of ≤10000 days between publication and retraction (32 papers) and bins with ≥5 papers (104 papers), leading to 1182 of 1318 papers (90%) plotted.

Confidence intervals capped at 100 for display.
